# Comparative analysis of capture methods for genomic profiling of circulating tumor cells in colorectal cancer

**DOI:** 10.1101/2022.06.02.22275905

**Authors:** Joao M. Alves, Nuria Estévez-Gómez, Monica Valecha, Sonia Prado-López, Laura Tomás, Pilar Alvariño, Roberto Piñeiro, Laura Muinelo-Romay, Patricia Mondelo-Macía, Mercedes Salgado, Agueda Iglesias-Gómez, Laura Codesido-Prada, Joaquin Cubiella, David Posada

**Affiliations:** CINBIO, Universidade de Vigo, 36310 Vigo, Spain; Galicia Sur Health Research Institute (IIS Galicia Sur), SERGAS-UVIGO; Institute of Solid state Electronics, Technische Universität Wien, Austria; Roche-Chus Joint Unit, Translational Medical Oncology Group, Oncomet, Health Research Institute of Santiago de Compostela (IDIS), Santiago de Compostela, Spain; Centro de Investigación Biomédica en Red de Cáncer (CIBERONC), Madrid, Spain; Liquid Biopsy Analysis Unit, Translational Medical Oncology Group, Health Research Institute of Santiago de Santiago de Compostela (IDIS), Santiago de Compostela, Spain; Department of Oncology, Hospital Universitario de Ourense, Research Group in Gastrointestinal Oncology-Ourense, Ourense, Spain; Department of Gastroenterology Hospital Universitario de Ourense, Research Group in Gastrointestinal Oncology-Ourense, Centro de Investigación Biomédica en Red de Enfermedades Hepáticas y Digestivas (CIBERehd), Ourense, Spain; Department of Biochemistry, Genetics, and Immunology, Universidade de Vigo, 36310 Vigo, Spain

**Author notes:** **Corresponding authors:** Joao M. Alves, David Posada. **Competing interests:** The authors declare no competing interests.

**Keywords:** circulating tumor cells, liquid biopsy, intratumor genomic heterogeneity, colorectal cancer, precision medicine

## Abstract

The genomic profiling of circulating tumor cells (CTCs) in the bloodstream should provide clinically relevant information on therapeutic efficacy and help predict cancer survival. However, the molecular characterization of CTCs has so far proven extremely difficult. A variety of technologies have been developed for CTC isolation, but so far the impact on the genomic assessment of CTCs has not been fully evaluated. To fill this gap, here we contrasted the genomic profiles of CTC pools recovered from blood samples obtained from four metastatic colorectal cancer (mCRC) patients using three different enrichment strategies (CellSearch, Parsortix, and FACS). Our results suggest clear differences in the mutational burden of CTC pools depending on the enrichment method used, with all evaluated methods returning a somewhat limited representation of the mutational spectrum of individual tumors, potentially due to allelic dropout during whole-genome amplification. Nevertheless, the CTC pools from Parsortix, and in part, CellSearch, showed diversity estimates, mutational signatures and drug-suitability scores remarkably close to the ones found in matching primary tumor samples. In contrast, FACS CTC pools were substantially enriched in apparent sequencing artifacts, which led to much higher estimates of genomic diversity. Although CTC genomics still faces technical challenges, our results suggest that CTC-derived metrics can reflect the diversity scores seen in primary tumor lesions thus highlighting the utility of CTCs to assess the heterogeneity status of individual tumors, and to help clinicians prioritize drugs in mCRC.

## 1. Introduction

Although research on cancer biology has traditionally been hampered by sampling issues, with most approaches relying on highly-invasive, risky, and, sometimes, difficult to obtain solid tissue biopsies (Marrugo-Ramírez, Mir, and Samitier 2018; Robertson and Baxter 2011; Chi 2016), strong evidence has emerged in recent years that the peripheral blood, as well as other body fluids, offer a valuable source of cancer-associated materials (K. Pantel and Speicher 2016; Crowley et al. 2013). As opposed to tissue biopsies, liquid biopsies represent a minimally invasive alternative to capture clinically-relevant information about tumors (Krebs et al. 2014), including circulating tumor cells (CTCs). CTCs are thought to consist of cells shed by the primary tumor (PT) and metastatic lesions into the bloodstream, and have become the subject of intense research due to their likely role in the metastatic process (Klaus Pantel and Alix-Panabières 2017; Krebs et al. 2014; Castro-Giner and Aceto 2020). In recent years, multiple studies have demonstrated the clinical significance of CTCs for prognosis and therapeutic management, with CTC burden being correlated with unfavorable overall survival in several cancer types (Magbanua et al. 2019; Silveira et al. 2021; Basso et al. 2021; Chemi et al. 2019).

Importantly, sequencing studies exploring the genomic landscape of CTCs (Chemi et al. 2019; Gulbahce et al. 2017; Magbanua et al. 2018; Court et al. 2020) have shown that the mutational profiles of CTCs generally reflects the overall genomic composition of matched primary and metastatic lesions. Collectively, these results suggest that CTC diversity represents the overall tumor heterogeneity probably better than single tumor tissue biopsies, and incorporating CTC genomic information is expected to increase the clinical value of liquid biopsies by providing better predictions of therapeutic sensitivity and survival outcome (Pailler et al. 2019; Paoletti et al. 2018).

Nevertheless, implementing a comprehensive molecular characterization of CTCs into routine clinical procedures has, so far, proven extremely challenging (Rossi and Zamarchi 2019; Kowalik, Kowalewska, and Góźdź 2017). Indeed, despite the numerous strategies already available for isolating CTCs (Bankó et al. 2019), ranging from methods based on physical properties of cells (i.e., size and deformability) to others based on biological characteristics (e.g., cell surface marker expression), CTCs are typically present at very low numbers in the blood and can show a wide range of phenotypes (Neves et al. 2021; Kowalik, Kowalewska, and Góźdź 2017; Hyun et al. 2016). As a consequence, these technologies are likely to differ in their detection sensitivity and recovery rates but, to date, comparative analyses across distinct technologies are lacking (Neves et al. 2021). Of particular interest to us, it remains unclear how the different isolation methods can have an impact on the assessment of the genomic landscape of CTCs.

In order to identify an efficient strategy for the downstream genomic profiling of CTCs in metastatic colorectal cancer (mCRC), here we contrasted three different CTC-enrichment approaches —CellSearch®, Parsortix® and Fluorescence-Activated Cell Sorting (FACS). While all methods evaluated struggled here with data quality issues, our results indicate that Parsortix and, in part, CellSearch are able to provide genomic heterogeneity scores, mutational signature profiles and therapeutic targets compatible with those found in matching primary tumor samples.

## 2. Material & Methods

### 2.1 Patient selection and blood collection

We enrolled four mCRC patients diagnosed between October 2017 and September 2019 at the Hospital Universitario de Ourense, Spain, with histologically proven CRC, and either therapy naive or showing evidence of progression. On the same day, we collected three blood samples per patient and stored them into three different containers, one for each CTC-enrichment protocol, at room temperature: CellSave Preservative tubes (Menarini Silicon Biosystems, Italy) for CellSearch, Transfix CTC-TVT tubes (Cytomark, UK) for Parsortix and cell-free DNA BCT CE tubes (Streck, NE, USA) for FACS. In addition, we obtained a formalin-fixed paraffin-embedded (FFPE) block of the primary tumor (PT) from each patient. Importantly, all specimens were obtained and collected after written informed consent from all subjects using a protocol approved by the Clinical Ethic Committee of Pontevedra-Vigo-Ourense (2018/301 approved 19/06/2018).

### 2.2 CTC enrichment

We processed all samples within 96 hours after the blood was drawn using three different strategies for CTC enrichment. The CellSearch® system (Menarini, Silicon Biosystems, Bologna, Italy) enumerates and isolates CTCs of epithelial origin (CD45-, EpCAM+, and CK8+, 18+, and/or 19+). The Parsortix® platform (ANGLE plc, UK) traps CTCs due to their larger size and lower compressibility than blood cells. The FACS strategy separates CTCs based on custom markers; in our case EpCAM+/CD45-/CK7,8+.

#### 2.2.1 CellSearch

For each sample, we processed 7.5 mL of whole blood in the CellTracks Autoprep system using the Circulating Tumor Cell Kit (Menarini, Silicon Biosystems, Bologna, Italy). This kit consists of ferrofluids coated with epithelial cell-specific anti-EpCAM antibodies to immuno-magnetically enrich epithelial cells and a mixture of antibodies directed to cytokeratins (CKs) 8, 18, and 19 conjugated to phycoerythrin (PE); an antibody to CD45 conjugated to allophycocyanin (APC); and a nuclear dye 4′,6-diamidino-2-phenylindole (DAPI). Afterward, we analyzed the processed samples with the CellTracks Analyzer II according to the manufacturer’s instructions. We identified the CTCs as round or oval cells with an intact nucleus (DAPI positive), CK positive and CD45 negative (**Fig. S1**). We stored the CTC-enriched samples at −80 °C.

#### 2.2.2 Parsortix

We loaded 7.5 mL of peripheral whole blood per sample into a Parsortix microfluidic device (Angle plc, UK). We enriched the samples in disposable Parsortix cassettes with a gap size of 6.5 µm (GEN3D6.5, Angle Inc., Guildford, UK) and at 99 mbar of pressure, according to the manufacturer’s guidelines. After separation, we collected the captured cells in 200 µL of PBS and stored them at −80 °C.

#### 2.2.3 Fluorescence-activated Cell Sorting (FACS)

In order to obtain the peripheral blood mononuclear cell (PBMC) fraction –to be used as healthy controls– we took 1 mL from each blood sample and performed Ficoll-Paque gradient centrifugation. We kept the PBMCs in RNA later (Ambion, TX, USA) at −80°C until genomic DNA (gDNA) extraction. The remaining blood volume (7-9 mL) was then used for CTC staining and collection. After validating our FACS protocol using spike-in experiments (see **Supplementary note 1**), we followed a similar approach to Miller et al. (2012) and first lysed the red blood cells using BD Pharm Lyse lysing solution (BD Biosciences, NJ, USA) following the fabricant recommendations. When needed, we repeated the lysing step up to three times. We then resuspended the cells in phosphate-buffered saline (PBS) solution and filtered them with a 70 μm cell strainer (Falcon, NY, USA). We used the FIX & PERM™ Cell Permeabilization Kit (Invitrogen, MA, USA) and incubated the filtered cell suspensions with antibodies (BD Biosciences, NJ, USA) against the epithelial cell adhesion molecule (EpCAM; PerCP-Cy5.5, IgG1λ, clone EBA-1) and the leukocyte common antigen CD45 (FITC, IgG1κ, clone HI30) with reagent A for 25 minutes in the dark at room temperature for fixation. Cells were then washed once with 500 µL of PBS and centrifuged at 200 x g for 5 minutes. We resuspended the cell pellet in 1 ml of PBS and incubated it with an antibody against the epithelial markers cytokeratins 7 and 8 (CK7,8; PE, IgG2ɑ/κ, clone CAM 5.2) and reagent B for 20 minutes in the dark at room temperature for permeabilization. We washed once again and resuspended the cells in 500 µl of PBS. Finally, we selected and collected 3 µl of PBS pools of CTCs based on an EpCAM+/CD45-/CK7,8+ phenotype (**Fig. S2**) using a FACSAria III (BD Biosciences, NJ, USA). We analyzed the data using the FACSDiva (BD Biosciences, NJ, USA) and FlowLogic software (Miltenyi Biotec, Germany).

### 2.3 Whole-genome amplification of CTC-pools

Given the large collection volume (∼200 µl) from both CellSearch and Parsortix, we initially performed genomic DNA (gDNA) extraction of the CTC enriched samples obtained from these platforms using the QIAamp DNA Blood Mini Kit (Qiagen, Germany) before performing whole-genome amplification (WGA) using the Ampli1 kit (Menarini Silicon Biosystems, Italy). For CellSearch and Parsortix samples, we carried out the WGA starting with 1 µl of DNA and CTC pools from FACS were amplified directly. In order to avoid contamination, we worked in a laminar-flow hood and used a dedicated set of pipettes and UV-irradiated plastic materials. We included positive (10 ng/µl REPLIg human control kit, Qiagen, Germany) and negative (DNase/RNase free water) controls during the amplification and used the Ampli1 QC Kit to evaluate the amplification. Samples with a positive signal for at least two PCR fragments were selected to increase the total dsDNA content using the Ampli1 ReAmp/ds kit. We then removed the kit adaptors by incubating at 37 °C for 3 h a mixture of 5 µl of NEBuffer 4 10X (New England Biolabs, MA, USA), 1 µl of MseI 50U/µl (New England Biolabs, MA, USA), 19 µl of nuclease-free water and 25 µl of dsDNA followed by a step at 65 °C for 20 min for enzyme inactivation. Finally, we purified the samples with 1.8X AMPure XP beads (Agencourt, Beckman Coulter, CA, USA), quantified the DNA yield with Qubit 3.0 fluorometer (Thermo Fisher Scientific, MA, USA) and checked the amplicon size distribution with the D1000 ScreenTape System in a 2200 TapeStation platform (Agilent Technologies, CA, USA).

### 2.4 FFPE and PBMCs bulk gDNA isolation

We performed the extraction of bulk gDNA from FFPE samples using the QIAamp DNA FFPE tissue kit (Qiagen, Germany) by incubating and shaking at 60 °C for 1 h before slicing them and adding a deparaffinization solution (DS). We performed a 56°C incubation step for 1 h and when needed, performed a second addition of DS in a new tube in order to remove remaining paraffin. We used the QIAamp DNA Blood Mini Kit (Qiagen, Germany) for the extraction of gDNA from PBMCs bulks and estimated DNA yield using the Qubit 3.0 fluorometer (Thermo Fisher Scientific, MA, USA) and DNA integrity with the Genomic DNA ScreenTape Assay (Agilent Technologies, CA, USA).

### 2.5 Whole-exome sequencing

CTC pools and bulk sequencing libraries were constructed at the Spanish National Center for Genomic Analysis (CNAG; http://www.cnag.crg.eu) with the SureSelect XT and Agilent Human Exon v5 kits (Agilent Technologies, CA, USA). In total, seven whole-genome amplified CTC-pools (two from FACS: P4 and P5, one from CellSearch: P1 and 4 from Parsortix: P1, P3, P4 and P5) and four FFPE bulk samples were sequenced at 100X and four PBMCs samples at 60X. All samples were run on an Illumina NovaSeq 6000 (PE100) at CNAG.

### 2.6 Data processing and variant calling

After trimming amplification and sequencing adapters from the raw FASTQ files, we aligned the sequencing reads from CTC pools, tumor and healthy samples to the Genome Reference Consortium Human Build 37 (GRCh37) using the MEM algorithm in the BWA software (Li 2013). Following a standardized best-practices pipeline (Van der Auwera et al. 2013), we filtered out reads with low mapping quality. We next performed a local realignment around indels, and removed PCR duplicates. We identified somatic single nucleotide variants (SNVs) for each CTC-capture method using the multi-sample variant-calling feature implemented in MuTect2 software, taking as input the BAM files of the different sample types available (i.e., tumor bulk + CTC-pool + healthy control). We then used FilterMutectCalls to remove calls in any sequence context artifacts or contamination fractions (but see **Supplementary note 2** and **Fig. S3**). Afterwards, the genotypes of variants showing a coverage depth ≥ 10, alternative allelic depth ≥ 2 and allele frequency estimates ranging from 0.05 to 0.75 were kept for downstream analysis. For all datasets, we merged the inferrred SNV calls and performed variant annotation using Annovar software (v.20200608) (Wang, Li, and Hakonarson 2010).

### 2.7 Mutational Signatures

For all datasets, we ran sigProfilerExtractor (Ashiqul Islam et al. 2021) under default parameters to identify *de novo* mutational signatures for single-base substitutions (SBS), followed by the assignment of the decomposed signatures to known COSMICv3 SBS96 signatures (Alexandrov et al. 2020).

### 2.8 PanDrugs

We used PanDrugs (Piñeiro-Yáñez et al. 2018) (http://www.pandrugs.org) –a web-based platform that attempts to match genomic data to available drug therapies in order to guide personalized treatment selection– to explore changes in therapeutic options and drug suitability scores across the different datasets. For that purpose, we first ran PanDrugs using a VCF with the list of exonic mutations identified in the tumor bulk sample of each patient to identify CRC-specific therapeutic candidates. Next, we extracted the drug score (that ranges from −1 to 1 and measures the suitability of each drug using a database of curated gene-drug relationships and the collective gene impact) to identify the top 25 therapeutic candidates in the PT samples. Afterwards, we performed a new query with PanDrugs using the exonic mutations in each of the CTC samples to examine whether the CTC-derived genomic information identifies similar therapeutic options and drug sensitivity scores as for the bulk.

### 2.9 MATH scores

For the datasets with an unknown number of CTCs or with CTC counts > 1, we additionally estimated the mutant-allele tumor heterogeneity (MATH) score (Mroz and Rocco 2013). The MATH score is based on the distribution of allele fractions among somatic mutations and is calculated as the percentage ratio of the width of the data to the center of its distribution:

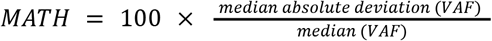

Importantly, since MATH scores are sensitive to unreliable allele frequency estimates stemming from sites with poor sequencing coverage depth, for their calculation we applied an additional filter to restrict our mutation calls to positions showing a depth of coverage ≥ 25 and alternative allelic depth ≥ 5.

## 3. Results

### 3.1 CTC-counts

Patient-level CTC counts were available for two of the selected CTC-capture methods (as the Parsortix platform does not provide CTC counts). Using the CellSearch system, we isolated one CTC from patient P1, whereas no CTCs were detected for the remaining patients. Importantly, although we were able to recover potential CTCs from all patients using FACS (P1 = 2 CTCs; P3 = 6 CTCs; P4 = 2 CTCs; P5 = 1 CTC), after whole-genome amplification we only obtained high-quality sequencing libraries for patients P4 and P5.

### 3.2 Tumor mutational burden

Across all patients, we found sharp differences in the number of somatic mutations (SNVs) identified with the different CTC-capture methods (**Fig. 1**). While the number of mutations called with both CellSearch and Parsortix datasets were close to, or lower than, the number of mutations observed in the matched PT bulk samples, the mutation counts in the FACS CTC pools were consistently much higher (by one order of magnitude) than in the bulk samples. Remarkably, regardless of the CTC-capture strategy, we found a very small overlap in mutation calls between CTC pools and PT datasets: Parsortix (average of 4.6%), CellSearch (3.4%) and FACS (0.4%).

**Figure 1.**
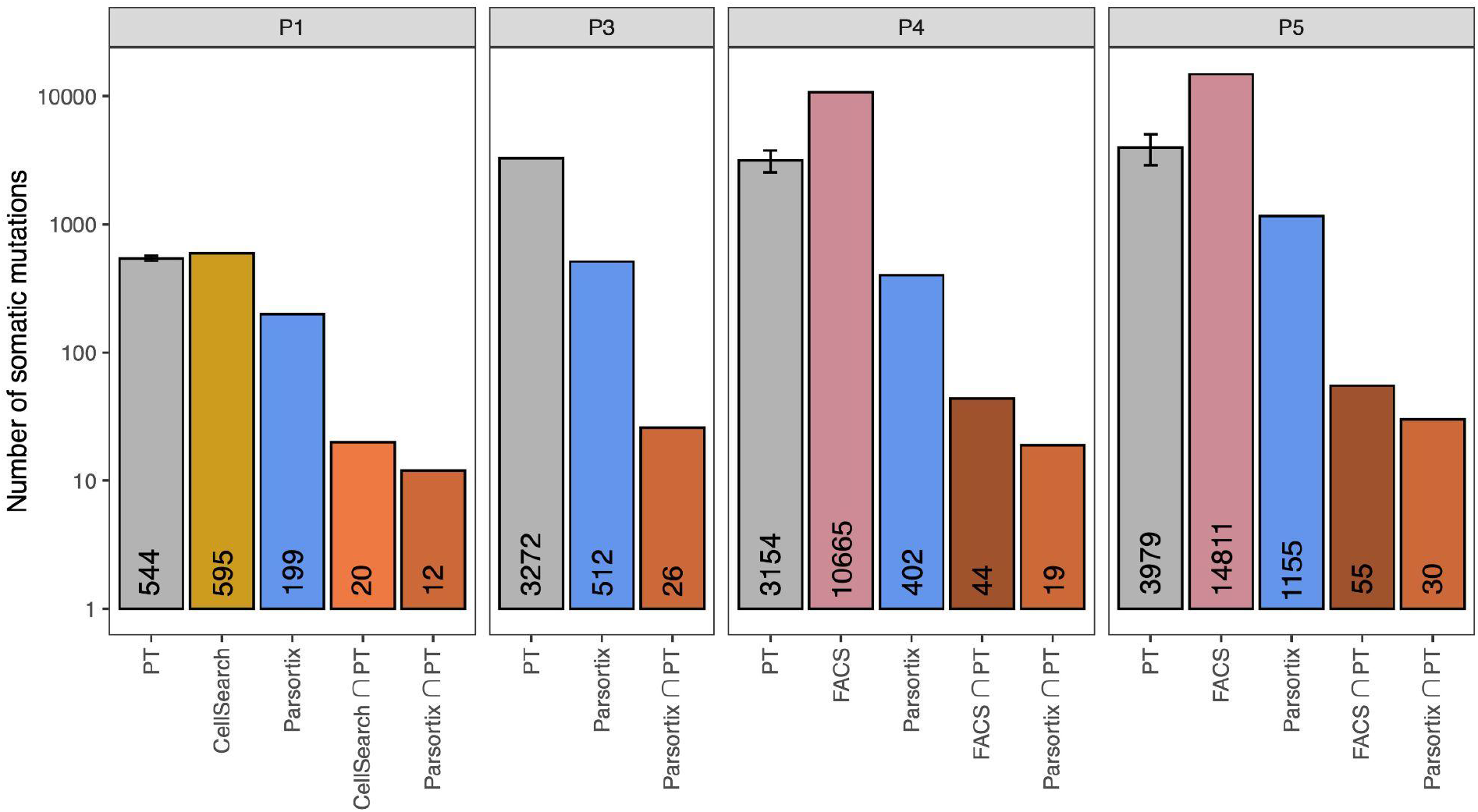
SNV abundance per CTC-capturing method. Barplots depicting the total number of SNVs identified by MuTect2 for each dataset. Number of SNVs shown at the bottom of each bar. Bar colors reflect the different input material or capturing method: gray = FFPE primary tumor (PT) sample; gold = CellSearch CTC pool; blue = Parsortix CTC pool; pink = FACS CTC pool. Error bars are only available for bulk tumor samples and reflect the 95% confidence interval. Orange bars depict the amount of shared sites between CTC-datasets and PT samples. The y-axis is in log scale.

Similarly, within each patient, the number of mutations shared between CTC pools captured with distinct methods was generally small (**Fig. 2a**). In any case, we identified a considerable number of CTC-specific mutations across datasets, including non-silent ones (**Fig. 2a** and **Fig. S4**).

**Figure 2.**
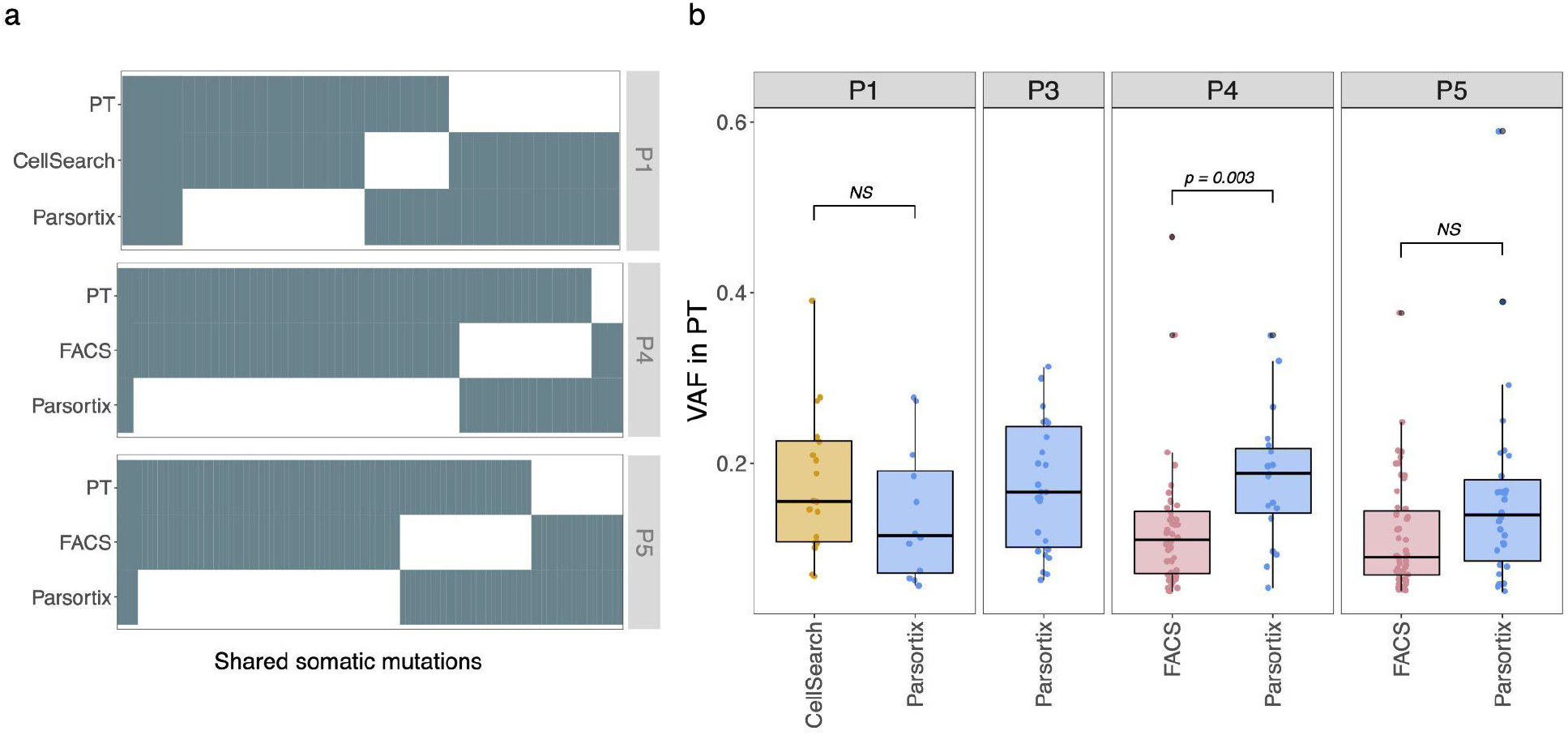
Genomic profiling and clonality of CTC-pools. **a**. Occupancy matrix of shared sites (P1 = 41 SNVs; P4 = 65 SNVs; P5 = 100 SNVs) across the CTC-capture methods within patients. Different colored tiles reflect different mutation status: dark grey=mutation; white=reference/missing data. Patient ID shown on the right. **b**. Boxplots depicting the bulk-level VAF estimates of the SNVs shared with the CTC pools. Boxplot colors represent the different CTC-capture methods. Statistical analysis was performed using the two sample KS-test to compare bulk VAF estimates of shared sites between CTC-capture methods. Significant *p-values* shown above boxplots.

Furthermore, within the PT samples, the median variant allele frequency (VAF) of the shared mutations with CTC pools ranged from 0.19 (Parsortix - P4) to 0.09 (FACS - P5), suggesting that the isolated CTCs are derived from minor subclones within the PTs (**Fig. 2b**). Similar VAF scores were observed between the CellSearch and Parsortix CTCs. In P4 we found a significant difference between Parsortix and FACS (**Fig. 2b**), with the FACS CTCs being generally enriched with mutations at low frequency.

Importantly, as shown in **Fig. 2b**, the CTC pools showed a clear depletion of clonal (i.e., allele frequency ≥ 0.4) mutations observed in the PT samples. While these results may appear surprising - as, in theory, clonal mutations in the PT should appear in all CTCs sampled-, the failure to identify such mutations can be, in part, explained due to the limited coverage breadth of the CTCs. Indeed, across all samples, only a small fraction (5 to 39%) of regions harboring clonal variants in the PTs was covered with sequencing reads in the CTC pools (**Table SI**). Moreover, after looking at the coverage statistics of heterozygous single-nucleotide polymorphisms (SNPs) (**Fig. S5**), we additionally found strong evidence of allele dropout (ADO) taking place during WGA. Despite being particularly obvious in the FACS datasets -which showed an averaged ADO of 67% for the heterozygous sites called-, all CTC pools showed some degree of ADO (CellSearch ADO: 9% and Parsortix ADO: 9%) which very possibly interfered with the identification of clonal mutations.

### 3.3 Measuring intratumor genomic heterogeneity (ITH)

Afterwards, we explored potential differences in intratumor genomic heterogeneity (ITH) estimates between PT and CTC samples by examining the frequency distribution of somatic mutations across the different datasets. Importantly, since several of our CTC samples comprised only one cell, the subsequent analyses were limited to datasets with either an unknown number of CTCs or CTC counts > 1, and included the Parsortix CTCs from patients P1, P3, P4 and P5, and the FACS CTCs from patient P4.

As illustrated in **Fig. 3a-b** (and **Fig. S6a-b**), we found significant differences in the distribution of VAF estimates between PT and CTC pools. In patient P4, specifically, both FACS and Parsortix datasets showed an enrichment towards low frequency variants when compared to the corresponding PT sample. Moreover, a quantile-quantile (Q-Q) plot further revealed contrasting differences in the skewness of the VAF distributions stemming from the different CTC-capture methods (**Fig. 3b**). Indeed, while the Parsortix CTCs encompassed a larger number of mutations at intermediate frequencies when compared to the matched PT sample, the FACS CTCs showed a substantial depletion of this class of mutations.

**Figure 3.**
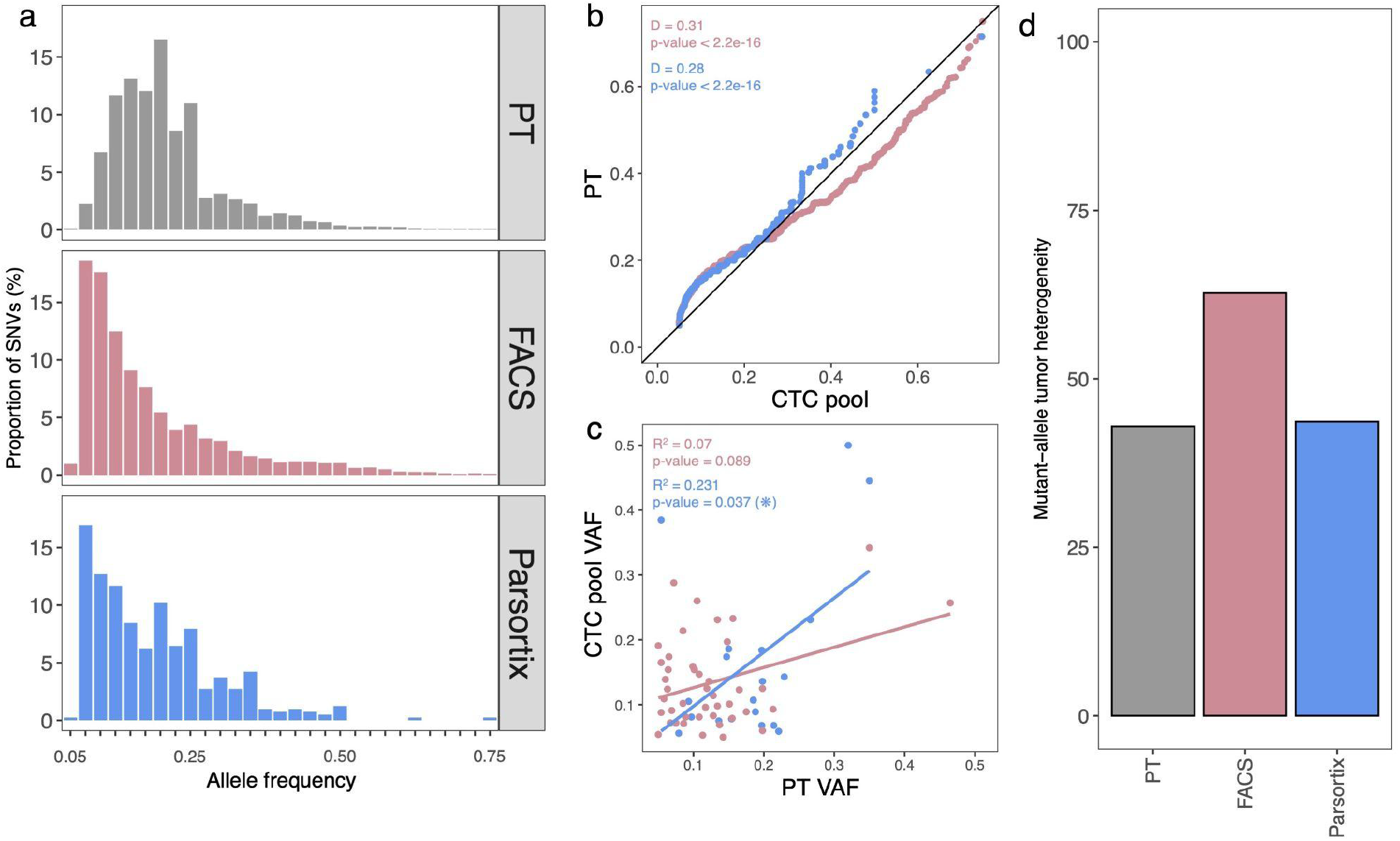
Measuring ITH through CTC pools. **a**. Histograms depicting the variant allele frequency (VAF) distribution of somatic mutations for the different datasets of patient P4. Different datasets highlighted with different colors: PT = grey; FACS = pink; Parsortix = blue with dataset ID shown on the right. Histograms scaled to percentages. **b**. Q-Q plot comparing the distribution of allele frequency estimates in CTC pools and PT samples of patient P4. Statistical analysis was performed using the two sample KS-test to compare the VAF distribution between CTC pools and PTsamples. KS D (distance) statistic and *p-values* are shown on the upper left side of the plot. **c**. Scatter plot describing the similarity of VAF scores of overlapping sites between CTC pools and PT samples. Solid lines represent the best fit from regression analysis. R^2^ scores and *p-values* are shown on the upper left side of the plot. **d**. Barplot depicting the MATH scores obtained using the mutation sets passing our strict filtering - Primary tumor = 416 SNVs; FACS = 8193 SNVs; Parsortix = 46 SNVs (see *methods*).

Interestingly, in the Parsortix CTCs, we found a significant positive correlation between the VAF estimates of shared mutations between the CTC pool and the PT sample (**Fig. 3c**). As for the FACS CTCs, although a positive trend was found, the relationship was not significant (similar to the results of the remaining datasets - **Fig. S6c**).

We also observed sharp differences in the levels of ITH, as measured by MATH scores, among CTC-capture methods. In patient P4, the Parsortix CTCs and the PT sample returned highly concordant MATH scores (43.6 and 43.0, respectively), while the FACS dataset displayed a much larger MATH score (62.8) (**Fig. 3d**). Importantly, very similar MATH scores were also observed between the remaining Parsortix CTCs and the PT samples (**Fig. S6d**).

### 3.4 Mutational signatures

We next explored the mutational signatures in the different datasets to look for potential differences between CTC pools and PT samples (**Fig. 4a**). Across all datasets, *SigProfilerExtractor* identified a total of five different mutational processes, with PT samples being predominantly enriched in “clock-like” (i.e., ageing) COSMIC signatures SBS1 and SBS5. A similar contribution of SBS1 and SBS5 was also found in the CellsSearch and Parsortix CTC datasets, albeit the lower cosine similarity score (cosine similarity: 0.505) obtained - likely due to the limited number of mutations available for signature assignment in these datasets. In contrast, FACS CTCs were predominantly characterized by signature SBS46 (65% in both cases), a mutational signature typically associated with sequencing artifacts.

**Figure 4.**
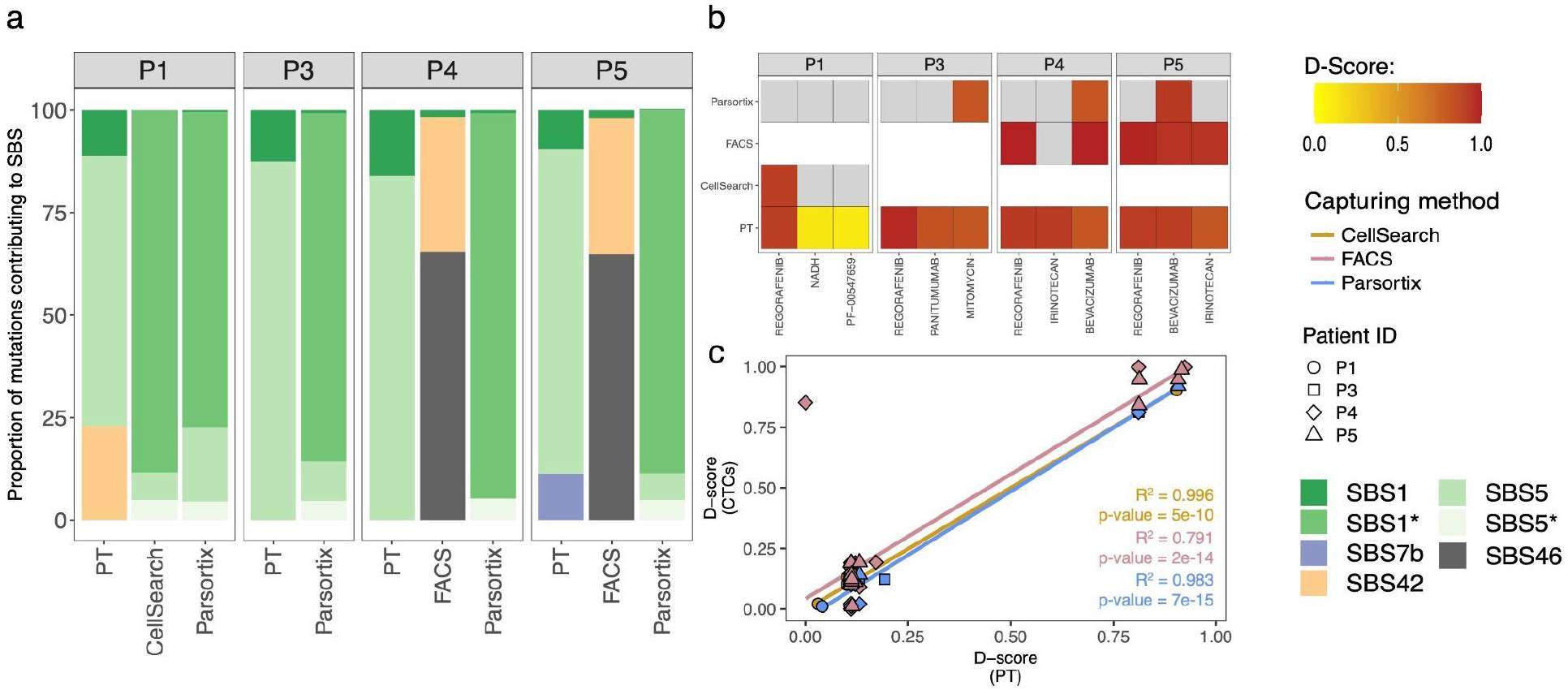
Mutational signatures and drug sensitivity concordance. **a**. Barplots depicting the proportion of mutations contributing to the different signatures/processes across the different samples. Sample IDs are shown at the bottom and patient IDs are shown at the top. Different colors reflect the identified mutational signature with the COSMIC SBS ID shown at the right of the plot. Legend asterisks distinguish mutational signatures identified by *SigProfilerExtractor* showing suboptimal cosine similarity scores (i.e., < 0.90). **b**. Tile plots depicting the overlap between the top-3 drug candidates in primary tumor (PT) samples (ordered from left to right) and corresponding CTC-pools. Tiles colored according to drug sensitivity score (D-score). Gray tiles correspond to therapeutic options not recovered by *PanDrugs* for that specific sample. **c**. Scatter plot depicting the correlation between the best therapeutic candidates (measured using the D-score) identified in both PT and CTC samples. Shape distinguishes the different patients while colors reflect the different CTC-capture methods. Solid lines represent the best fit from regression analysis. R^2^ scores and *p-values* are shown on the bottom right side of the plot.

### 3.5 Drug suitability scores

Finally, using the list of exonic positions available, we collected the best drug candidates for each dataset in order to evaluate potential changes in the type and sensitivity of therapeutic options between CTC pools and PT samples (**Fig. 4b**). Across patients, both FACS and CellSearch CTCs recovered, in most cases, the top candidate drugs (i.e., with the highest D-Score) identified with the corresponding PT samples. The Parsortix CTCs, on the other hand, often returned only a subset of treatment options identified in the matched PT. In any case, the fact that all CTC-capture methods showed a significant positive correlation in drug sensitivity scores between CTCs and PT samples (**Fig. 4c**) appears to indicate that the CTC genomic profiles offer reliable information for prioritizing therapeutic strategies in CRC.

## 4. Discussion

The isolation of CTCs remains challenging largely because of their scarcity in peripheral blood and phenotypic heterogeneity. While recent technological advances have resulted in improved capture strategies, most available methods differ in key aspects such as enrichment efficiency, cell viability, and throughput. Studies evaluating the implications of different CTC-capture methods on downstream analyses are scarce, and it remains unclear whether distinct enrichment approaches can provide compatible descriptions of the mutational landscape of CTCs.

In this study, in order to evaluate the impact of different CTC-capture strategies on the downstream molecular characterization of CTCs, we contrasted the genomic profiles of primary tumor samples against CTC pools recovered using three different enrichment strategies in four mCRC patients.

Our whole-exome sequencing experiments suggest differences in the mutational loads of CTC pools due to the enrichment method used. In sharp contrast to the results obtained with the CellSearch and Parsortix systems, the mutation counts in our FACS CTC pools exceeded by an order of magnitude the number of mutations observed in the corresponding primary tumor samples. Importantly, the FACS datasets showed significant enrichment for mutations at lower frequencies, with a high proportion of these mutations being later linked to a mutational signature associated with sequencing artifacts (i.e., SBS46).

These results suggest that our FACS-derived CTCs might have accumulated DNA lesions along with the different steps of the protocol. Unfortunately, identifying and subsequently removing these potential errors is not necessarily straightforward. Although one could argue that setting lower and upper bounds on the minor allele frequency could potentially prevent downstream variant call artifacts, it should be noted that sequencing CTC pools must necessarily be preceded by multiple rounds of genomic amplification. Since most WGA methods inevitably introduce biases in the resulting sequencing data (e.g., uneven genome amplification, allelic imbalances and dropout), for any given site, the derived allele frequency score will not necessarily reflect its true frequency in the CTC population sampled. Indeed, in our datasets, these biases were reflected both in the imbalanced distribution of allele frequencies of heterozygous SNPs and in the relatively poor concordance in VAFs of shared somatic sites between CTC pools and PT samples.

In any case, and for all datasets analyzed, Parsortix-derived CTC pools provided similar descriptions of ITH when compared to the corresponding PT samples, with MATH scores in clear agreement with previous estimates in mCRC (Bettoni et al. 2019). In contrast, this metric was largely overestimated in our FACS dataset, perhaps as a consequence of the massive amount of potential sequencing artifacts. Moreover, while the identification of therapeutic candidates was not always identical between CTC pools and PT samples, all CTC pools analyzed suggested drug-candidates displaying a significant probability of response, and highly concordant sensitivity scores with the PT samples, thus providing strong evidence that CTC-based mutational profiles may contribute with valuable guidance for refining treatment tailoring (Khoo et al. 2016; Siravegna et al. 2017; Parikh et al. 2019).

## 5. Conclusion

In conclusion, CTC genomics still faces technical challenges for straightforward clinical applications. As seen throughout our study, all the methods evaluated struggled with data quality issues - potentially caused by the inherent technical bias introduced by the limiting amounts of input material (Navin 2014) and/or background DNA contamination (e.g., white blood cells) (Xu et al. 2015) – which resulted in a somewhat incomplete picture of the mutational landscape of these tumors. Nevertheless, and despite the limited number of patients analyzed and the failure of some enrichment methods to recover CTCs for all patients, it is important to highlight that the CTC pools recovered from Parsortix and, in part, CellSearch returned comparable ITH estimates, similar mutational signature profiles, and suggested equivalent therapeutic targets, when compared to those found in matching primary tumor samples.

On this basis, as the performance of technologies for the detection and isolation of CTCs continues to improve, allowing for more accurate and informative genomic data to be produced (Diamantopoulou, Castro-Giner, and Aceto 2020), future studies (using larger cohorts) should explore whether the mutational landscape and genomic diversity of CTC populations can indeed provide clinically relevant prognostic and predictive information beyond simple enumeration.

## Data Availability

All raw whole exome sequencing data used in this study will be available at the Sequence Read Archive database.

## Declarations

### Ethics approval and consent to participate

All specimens used in this study were obtained and collected after written informed consent from all subjects using a protocol approved by the Clinical Ethic Committee of Pontevedra-Vigo-Ourense (2018/301 approved 19/06/2018).

### Availability of data and materials

We have deposited raw whole-exome sequencing data at the Sequence Read Archive database under the accession code XXXXXXX (to be provided).

### Authors’ contributions

D.P. and J.M.A. conceived the study and designed the analyses. M.S. and J.C. collected the blood and tumor samples and patient information. N.E.G., S.P.L. and P.A. processed the samples, performed the spike-in experiments, obtained the CTC pools through FACS, and performed whole-genome amplification. R.P, L.M.R and P.M collected the CTC pools from CellSearch and Parsortix. J.M.A., L.T., and M.V. performed the analyses. All authors read and approved the final manuscript.

## Acknowledgements

We would like to thank all members of the lab for their comments on earlier versions of the manuscript. We also thank the Supercomputation Center of Galicia (CESGA) for providing all computational resources and Mercedes Peleteiro, the flow cytometry core facility manager, for her help and support.

