## Supplementary material for "Comparative analysis of capture methods for genomic profiling of circulating tumor cells in colorectal cancer"

### *Additional file 1*

Supplementary note 1. FACS protocol validation using Spike-in experiments.

Supplementary note 2. Variant calling concordance in primary tumor datasets.

Table SI. Clonal mutations (in primary tumors) identified in CTC-pools.

Figure S1. FACS gating strategy.

Figure S2. Concordance between primary tumor callsets.

Figure S3. SNV abundance per mutation type for each CTC-capture method.

Figure S4. Allele frequency distribution of heterozygous SNPs for each CTC-capture method.

Figure S5. Genetic heterogeneity in Parsortix datasets across remaining patients.

**Supplementary note 1. FACS protocol validation using spike-in experiments.**

For the validation of the CTC-enrichment protocol using fluorescence-activated cell sorting (FACS), we performed a series of spike-in experiments using cells from the Caco-2 cell line (American Type Culture Collection; https://www.atcc.org) - a polyploid colorectal cancer cell line which expresses the epithelial markers EpCAM and cytokeratin (CK) 7,8 and can be used as a model for epithelial EpCAM+/CK 7,8+ CTCs (Costa et al. 2012). First, we grew Caco-2 in media consisting of Dulbecco's Modified Eagle's Medium/F12 with 3.151 g/l of glucose and L-glutamine (Lonza, Switzerland) and 1% penicillin/streptomycin (Lonza, Switzerland) at a working concentration of 100 units of potassium penicillin and 100 µg of streptomycin sulfate per 1 ml of culture media under an atmosphere containing 5 % $CO_2$ at 37 ºC. Once the cell culture was confluent, we harvested cells by incubating with accutase for 5 min at room temperature. Next, we washed cells with phosphate buffered saline (PBS) before staining and we incubated cells with antibodies (BD Biosciences, NJ, USA) against EpCAM, the leukocyte common antigen CD45 and CK 7,8. As cytokeratins correspond to intracellular markers, cells were fixed and permeabilized. To do this, we used the FIX & PERM™ Cell Permeabilization Kit (Invitrogen, MA, USA). After staining, we washed cells with PBS and immediately collected them on a FACS Aria III (BD Biosciences, NJ, USA). We used DRAQ5 to select nucleated cells and sorted double positive cells for EpCAM and CK 7,8 at doses of 5, 10, 25 and 50 cells. Next, a peripheral blood sample (8 mL) from a healthy donor was collected into heparinized tubes (BD Biosciences, NJ, USA) and we split it into 4 aliquots of 2 ml. Then, we added the isolated CTCs to the aliquots at the doses cited above and processed the blood by lysing the red blood cells with the BD Pharm Lyse lysing solution (BD Biosciences, NJ, USA) following the fabricant recommendations. We washed and prepared cell suspensions in PBS and acquired the double-stained EpCAM+/CK7,8+ cells using the FACS Aria III (BD Biosciences, NJ, USA) cytometer. We retrieved 2/5, 3/10, 15/25 and 35/50 double positive cells thus indicating a recovery efficiency of 40, 30, 60 and 70 %, respectively, for each condition.

**Supplementary note 2. Variant calling concordance in primary tumor datasets.**
Since the main goal of this study was to compare the genomic profiles of CTCs stemming from different capture technologies, we performed mutation calling by running MuTect2 independently for each CTC-capture method, rather than using a joint calling with all samples at our disposal (i.e., combining BAM files from different CTC-pools). An example command-line is shown below:

```
$ MuTect2 \
      -R [ReferenceGenome.fa] \
      -I PBMCs.P1 \
      -I PT.Bulk.P1 \
      -I Pool.Parsortix.P1 \
      -normal PBMCs.P1 \
      -[additional parameters] \
      -O P1-Parsortix-MuTect2.vcf
```

The rationale behind our approach was primarily to (1) approximate our calling strategy to a realistic scenario (as, in theory, CTC-based experiments generally rely on a single capturing method) and (2) avoid the expected increase in statistical power towards low frequency variants which could very possibly enrich our callsets in mutations that would otherwise be ignored during variant calling. As a consequence, with the exception of patient P3, multiple primary tumor callsets were generated for each patient. Throughout our study, all comparative analysis between primary tumor and CTC samples were performed by contrasting callsets obtained from the same runs (taking the example above, the Parsortix CTCs calls from patient P1 were compared against primary tumor mutations identified in the same MuTect2 run). In **Fig. S3** we show that, despite some slight differences, the variant sites in primary tumor callsets are extremely consistent across MuTect2 runs, which indicates that the impact of these minor differences is likely not significant.

**Table SI. Clonal mutations in the primary tumor samples (PT) identified in the CTC-pools.** Proportion of mutations shown in brackets. We considered clonal mutations those with allele frequency ≥ 0.4.

| Patient | Sample | Clonal mutations in PT | Clonal mutations in PT Covered in CTC pool ( ≥ 10 reads) | Clonal mutations in PT Mutated allele found in CTC pool |
|---|---|---|---|---|
| P1 | CellSearch | 20 | 2 (0.10) | 0 (0.00) |
| P1 | Parsortix | 20 | 1 (0.05) | 0 (0.00) |
| P3 | Parsortix | 60 | 13 (0.22) | 2 (0.03) |
| P4 | FACS | 164 | 33 (0.20) | 2 (0.01) |
| P4 | Parsortix | 164 | 50 (0.30) | 0 (0.00) |
| P5 | FACS | 61 | 15 (0.25) | 1 (0.03) |
| P5 | Parsortix | 61 | 24 (0.39) | 1 (0.03) |

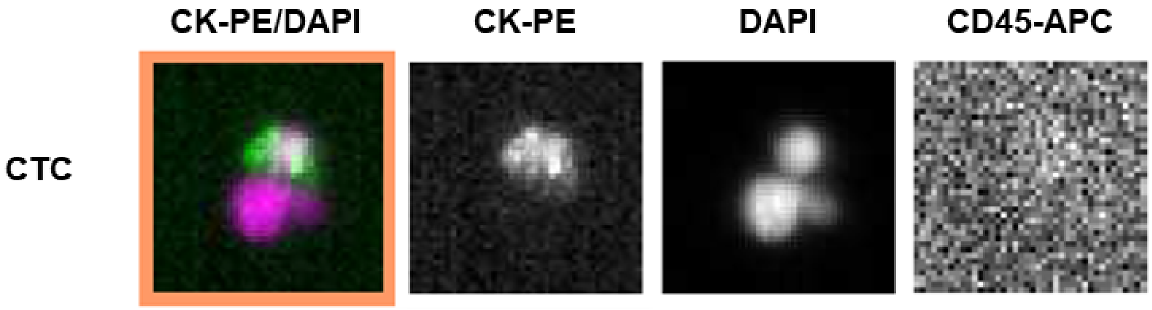


**Figure S1. CTCs captured by the CellSearch® system.** Images of the CTC isolated from the peripheral blood of patient P1 captured by the CellSearch® system. CTC is identified by round-oval morphology, positive staining for cytokeratins 8, 18, and 19 (CK-PE, phycoerythrin-conjugated antibody) and DAPI (DNA dye), and negative staining for CD45 (CD45-APC, allophycocyanin-conjugated antibody). In the left image, the CTC is visualized next to a nucleus (magenta).

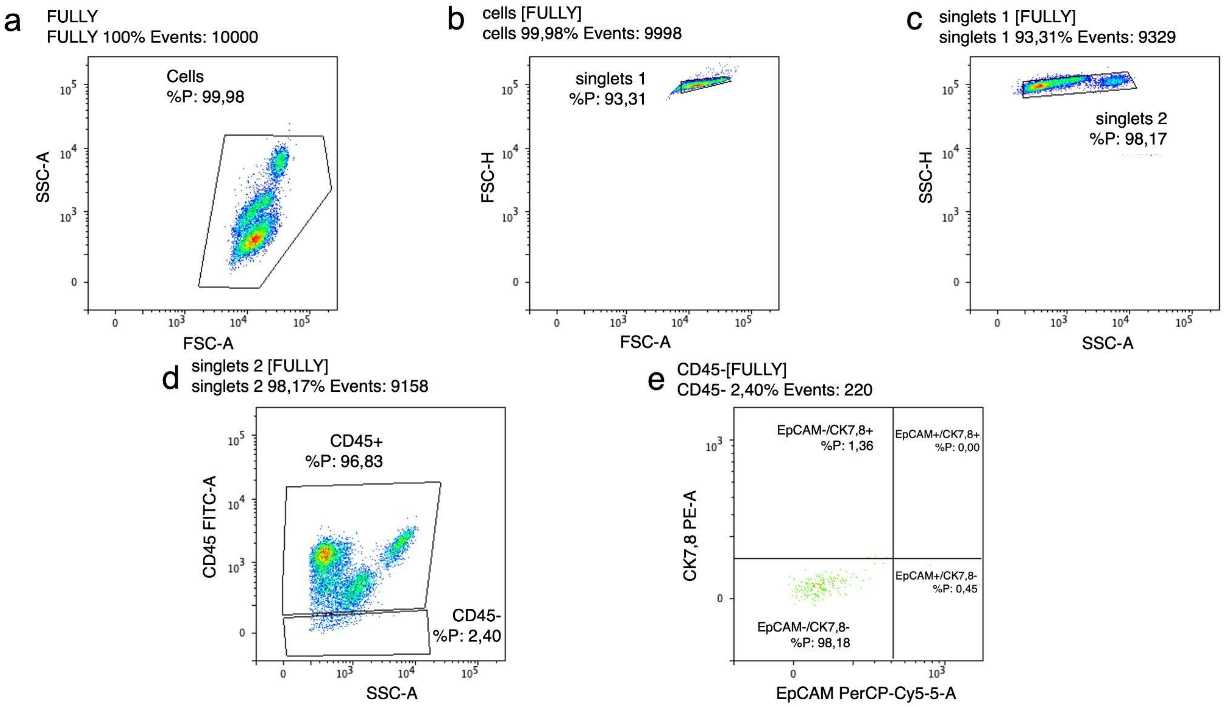


**Figure S2. FACS gating strategy for CTC-pools collection.** Representative pictures of sorted cells from patient P5. **a.** We used FSC/SSC plot to exclude cell debris. **b-c.** We then removed doublets and triplets based on FSC/SSC scattering. **d.** We selected CD45- cells. **e.** Finally, using a two parameter density plot (EpCAM vs CK7,8), we sorted double positive cells (EpCAM+/CK7,8+).

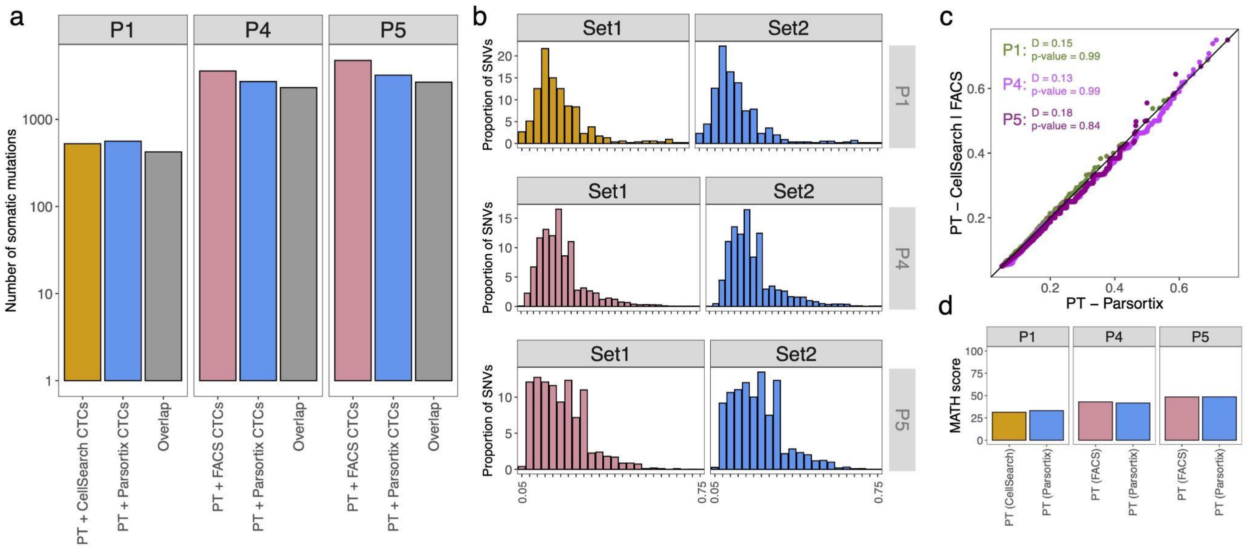


**Figure S3. Concordance between primary tumor mutations. a.** Barplots depicting the total number of (and the shared) SNVs identified in the primary tumor callsets. Color of the bars reflect the different MuTect2 runs. **b.** Histograms depicting the variant allele frequency (VAF) distribution of somatic mutations for the different primary tumor callsets (Patients P1, P4 and P5). Color of the bars reflect the different MuTect2 runs. **c.** Q-Q plot comparing the distribution of allele frequency estimates between primary tumor callsets. Different patients highlighted with different colors: P1 - dark green; P4 - orchid; P5 - purple. KS D (distance) statistic and p-values shown on the upper left side of the plot. **d.** Barplot depicting the MATH scores obtained using the variant sites passing our strict filtering for each primary tumor callset. Color of the bars reflect the different MuTect2 runs.

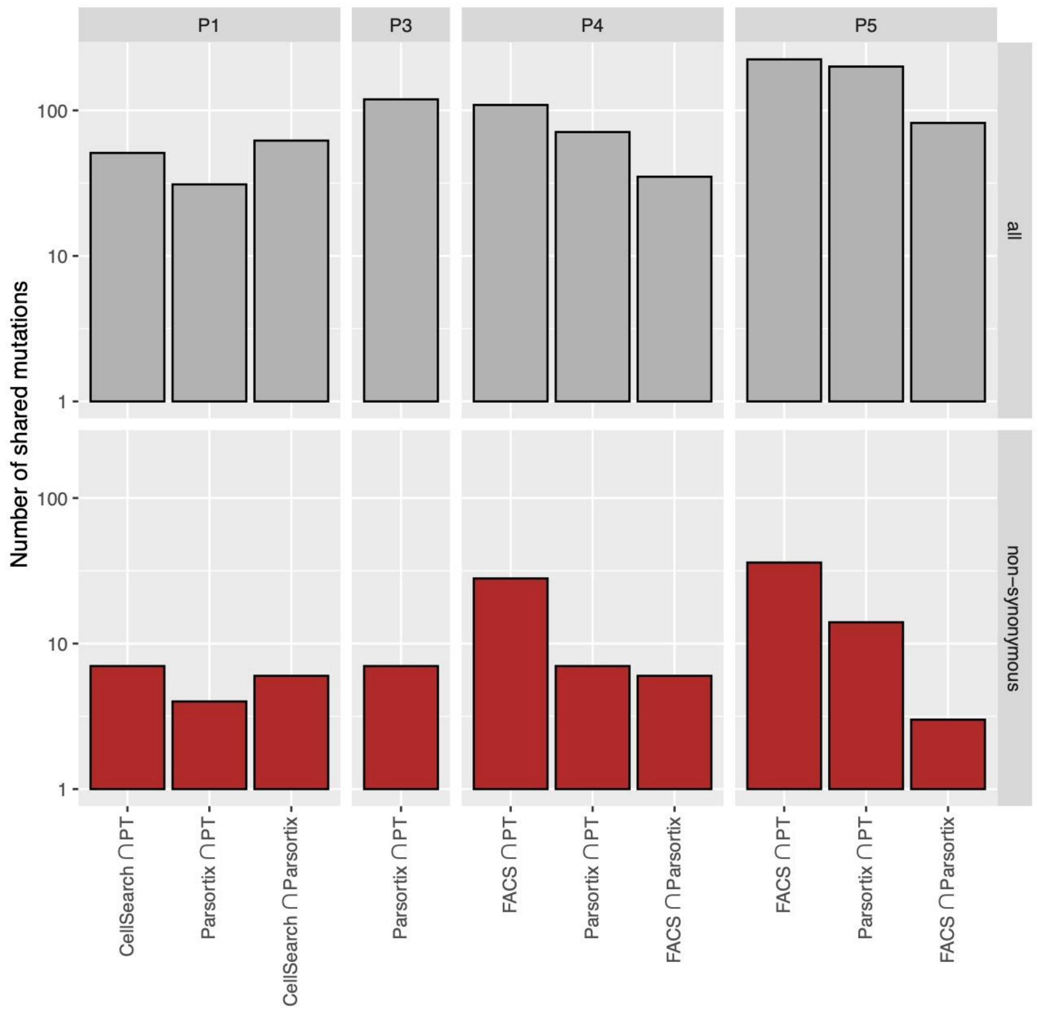


**Figure S4. SNV abundance per CTC-capture method.** Barplots depicting the number of shared SNVs identified by MuTect2 between CTCs and primary tumor samples. Color of the bars reflect the different types of somatic mutation: all = gray; non-synonymous = red.

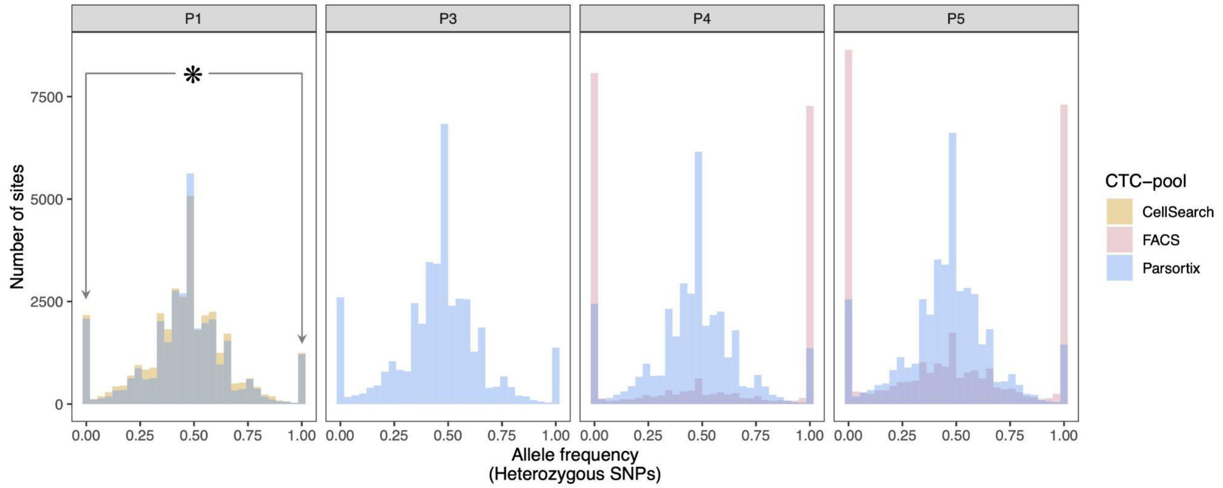


* Extreme allelic imbalance where only one of the alleles gets amplified and (subsequently) sequenced.

**Figure S5. Allele frequency distribution of heterozygous SNPs per CTC-capture method.** Histograms depicting the allele frequency distribution of heterozygous SNPs for each patient. The color of the histograms reflect different CTC-capture methods.

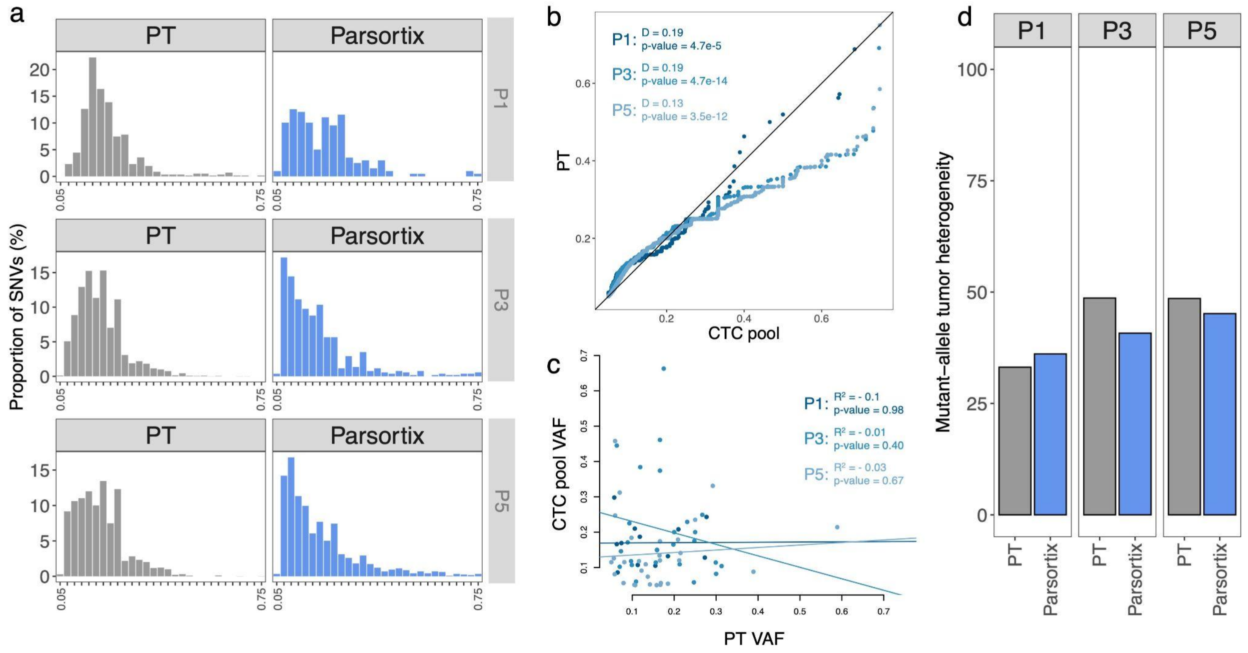


**Figure S6. Genetic heterogeneity in Parsortix datasets across remaining patients. a.** Histograms depicting the variant allele frequency (VAF) distribution of somatic mutations for the different Parsortix datasets (Patients P1, P3 and P5). **b.** Q-Q plot comparing the distribution of allele frequency estimates in CTC-pools and corresponding primary tumor samples. Different datasets highlighted with different shades of blue. Statistical analysis was performed using the two sample KS-test to compare the VAF distribution between CTC-pools and primary tumor samples. KS D (distance) statistic and p-values shown on the upper left side of the plot. **c.** Scatter plot describing the similarity of VAF scores of overlapping sites between CTC-pools and primary tumor samples (P1 = 12 SNVs, P3 = 26 SNVs, P5 = 30 SNVs). As in b., different datasets highlighted with different colors. Solid lines represent the best fit from regression analysis. R2 scores and p-values show on the upper left side of the plot. **d.** Barplot depicting the MATH scores obtained using the mutation sets passing our strict filtering - P1: PT = 135 SNVs & CTC pool = 30 SNVs; P3: PT = 731 SNVs & CTC pool = 90 SNVs; P5: PT = 717 SNVs & CTC pool = 315 SNVs (but see methods).
